# USING ARTIFICIAL INTELLIGENCE TO PREDICT TREATMENT OUTCOMES IN PATIENTS WITH NEUROGENIC OVERACTIVE BLADDER AND MULTIPLE SCLEROSIS

**DOI:** 10.1101/2025.06.17.25329802

**Authors:** Oliva H. Chang, Jaylen Lee, Felicia Lane, Michael Demetriou, Peter D. Chang

## Abstract

**Introduction and Objectives:** Many women with multiple sclerosis (MS) experience neurogenic overactive bladder (NOAB) characterized by urinary frequency, urinary urgency and urgency incontinence. The objective of the study was to create machine learning (ML) models utilizing clinical and imaging data to predict NOAB treatment success stratified by treatment type.

**Methods:** This was a retrospective cohort study of female patients with diagnosis of NOAB and MS seen at a tertiary academic center from 2017-2022. Clinical and imaging data were extracted. Three types of NOAB treatment options evaluated included behavioral therapy, medication therapy and minimally invasive therapies. The primary outcome – treatment success was defined as > 50% reduction in urinary frequency, urinary urgency or a subjective perception of treatment success. For the construction of the logistic regression ML models, bivariate analyses were performed with backward selection of variables with p-values of < 0.10 and clinically relevant variables applied. For ML, the cohort was split into a training dataset (70%) and a test dataset (30%). Area under the curve (AUC) scores are calculated to evaluate model performance.

**Results:** The 110 patients included had a mean age of patients were 59 years old (SD 14 years), with a predominantly White cohort (91.8%), post-menopausal (68.2%). Patients were stratified by NOAB treatment therapy type received with 70 patients (63.6%) at behavioral therapy, 58 (52.7%) with medication therapy and 44 (40%) with minimally invasive therapies. On MRI brain imaging, 63.6% of patients had > 20 lesions though majority were not active lesions. The lesions were mostly located within the supratentorial (94.5%), infratentorial (68.2%) and 58.2 infratentorial brain (63.8%) as well as in the deep white matter (53.4%). For MRI spine imaging, most of the lesions were in the cervical spine (71.8%) followed by thoracic spine (43.7%) and lumbar spine (6.4%).10.3%). After feature selection, the top 10 highest ranking features were used to train complimentary LASSO-regularized logistic regression (LR) and extreme gradient-boosted tree (XGB) models. The top-performing LR models for predicting response to behavioral, medication, and minimally invasive therapies yielded AUC values of 0.74, 0.76, and 0.83, respectively.

**Conclusions:** Using these top-ranked features, LR models achieved AUC values of 0.74-0.83 for prediction of treatment success based on individual factors. Further prospective evaluation is needed to better characterize and validate these identified associations.

## Introduction

Multiple sclerosis (MS) is a neuroinflammatory condition characterized by demyelination of the central nervous system.^1^ Roughly 1 million patients in the United States are affected by multiple sclerosis. It is the most common cause of disability in persons between the ages of 21 and 40 years old.^1,2^ Within 10 years of diagnosis, nearly every patient with MS will experience neurogenic lower urinary tract dysfunction.^2^ For 10% of patients, urinary complaints are the first sign of multiple sclerosis, ^2^ with more than half of MS patients would describe their urinary symptoms as moderate to severe.^3^

Neurogenic lower urinary tract dysfunction (NLUTD) is defined as the “abnormal function of either the bladder, bladder neck, and/or its sphincters related to a neurologic disorder.”^4^ Neurogenic bladder is another term that has been used for urinary dysfunction due to a neurologic condition. Broadly speaking, NLUTD is categorized as dysfunction with a) bladder storage or with bladder emptying or b) urethral sphincter contraction or relaxation. This results in a spectrum of urinary conditions including urinary frequency, urinary urgency, urinary incontinence, or urinary retention. NLUTD etiology in patients with multiple sclerosis is multifactorial with bladder spasticity and lesions along the spinal cord, thus contributing to a wide spectrum of urinary symptoms experienced by MS patients.

One type of NLUTD, neurogenic overactive bladder (NOAB), can significantly impact the quality of life especially with urinary incontinence (UI) and frequency, and has the potential for renal upper tract deterioration if presented with elevated bladder pressures. Patients with UI have a decreased health-related quality of life compared to those without UI.^5^ Treatment for NOAB includes behavioral therapies, medication therapies and minimally invasive therapies; however, treatment adherence rates are less than 40% at 1 year due to undesirable side effects and inadequate treatment outcomes.^6^ Currently, NOAB treatment options are similar between patients with idiopathic/non-neurogenic overactive bladder (OAB) or due to a neurogenic cause as in the case of MS.

The current paradigm for OAB or NOAB is not tailored to the individual and defaults to trialing various therapies. Therefore, the primary objective of this study is to utilize machine learning modeling to determine predictors for treatment success for neurogenic overactive bladder in patients with multiple sclerosis stratified by treatment type.

## Materials & methods

A retrospective cohort of patients were identified from a single academic tertiary medical center between 2017 and 2022. Inclusion criteria included female patients, 18 years or older, with diagnosis of multiple sclerosis and NLUTD. Only patients evaluated by both a urologist/urogynecologist and a neurologist were included. Additionally, only those with existing magnetic resonance imaging (MRI) brain and/or spine imaging with corresponding reports were included. Patients were excluded if imaging reports were not available or if urologic/neurologic care was established outside of the health system.

Clinical data, laboratory testing, and imaging data were manually extracted by two trained physicians. Additionally, variables related to MS were abstracted from clinical charts (duration and type of MS) as well as medication treatment including: capoxone, b-cell deplete, teriflunomide, fumarates, cladribine, alpha-4-integrin-blockers, Sphingosine-1-phosphate receptor modulators and immunomodulating medications (interferon).

Clinical data, testing and imaging reports for urologic and urogynecologic evaluations were reviewed. Baseline demographics, medical, surgical and gynecologic history was abstracted with specific focus on presence of pelvic organ prolapse, stress urinary incontinence, prior pelvic organ prolapse surgeries and recurrent urinary tract infections (UTIs). Voiding patterns such as day and nighttime frequency, urinary urgency, objective (measured by post-void residual) and subjective sensation of incomplete bladder emptying were recorded. For urodynamics, both the filling and voiding phase of the tests were recorded. Variables abstracted included bladder sensation, bladder capacity, bladder compliance, presence of urinary incontinence with/without abdominal and detrusor leak point pressure, detrusor overactivity. If fluoroscopic urodynamic was performed, variables such as bladder contour, bladder neck funneling and vesicoureteral efflux was recorded.

For each MRI exam, based on the radiology reports, MS lesions were characterized based on the location of lesions, number of lesions, activity of lesion, diffusion restriction and lesion enhancement. Additionally, the total brain volume loss was recorded. For each category, a standardized data collection template was developed in collaboration with an expert neuroradiologist.

Three types of NOAB treatment options were evaluated and categorized to include behavioral therapy, medication therapy, and minimally invasive therapies, such as percutaneous tibial nerve stimulation (PTNS), chemodenervation of bladder or sacral neuromodulation per American Urologic Association 2024 guidelines.^7^ The primary outcome, treatment success, was defined as > 50% reduction in urinary frequency, urinary urgency, or a subjective perception of treatment success as documented in clinical records.

Descriptive statistics were applied with continuous variables presented with means and standard deviation (SD) or medians and interquartile range (IQR).. Non-ordinal categorical variables were transformed into one-hot encoded vectors, thereby preventing the model from inferring spurious ordinal relationships. For N-class ordinal categorical variables, an integer encoding scheme was employed, mapping values to the range [0, N−1], preserving their inherent order. It is important to note that no data imputation techniques were required, as the dataset exhibited no missing values. Model-based and model-free techniques were designed for prediction of treatment success by treatment type. Feature selection aimed at identifying the most informative variables for model training, thereby reducing dimensionality and enhancing interpretability. Subsequently, feature rankings were derived using univariate statistical methods (t-test and chi-square test with bivariate analyses) to assess the individual predictive strength of each feature stratified by treatment type. For supervised learning, multiple machine learning algorithms were employed to build the classification model. For the model-based approach, the 10 most statistically and clinically relevant features for each treatment subtype were then fit into a LASSO (Least Absolute Shrinkage and Selection Operator) logistic regression (LR) machine learning model. Model-free techniques adapt to the dataframe without the use of a prior models with application of fewer assumptions. Model-free techniques employ non-parametric models with potential for ongoing retraining with the use of various methods such as XGBoost (Extreme Gradient Boosting)^8^ and SHAP plot after applying the top 10 features. Each model was trained and validated using the selected feature set to predict the target outcome. The choice of these diverse algorithms allowed for a comprehensive assessment of different modeling approaches and their respective strengths in handling the dataset’s characteristics.

For all modeling, the dataset was split into a 70% training and 30% validation cohort. Overall model performance was assessed using accuracy, sensitivity, specificity, PPV, NPV and receiver operating characteristic curves (ROC) from the corresponding area under the curve (AUC). Out of sample receiver operating characteristic curves (ROC) were generated for all models on the testing cohort with corresponding area under the curve (AUC). Analyses were conducted on R 4.2.2^9^ using the glmnet^10^, xgboost^11^, and SHAPforxgboost^12^ packages.

## Results

248 patients were screened and 110 patients met inclusion and exclusion criteria. The mean age of patients were 59 years old (SD 14 years), with a predominantly White cohort (91.8%), post-menopausal (68.2%). Patients were stratified by the NOAB treatment therapy type received with 70 patients (63.6%) at behavioral therapy, 58 (52.7%) with medication therapy and 44 (40%) with minimally invasive therapies. On MRI brain imaging, 63.6% of patients had > 20 lesions though majority were not active lesions. The lesions were mostly located supratentorial (94.5%), infratentorial (68.2%) brain and 58.2% in the deep white matter. For MRI spine imaging, most of the lesions were in the cervical spine (71.8%) followed by thoracic spine (43.7%) and lumbar spine (6.4%).

The entire cohort was then categorized by any use of behavioral, medication or minimally invasive therapies. Bivariate analyses between success and no success were performed stratified for each treatment strategies and those with a p value of < 0.10 was selected as features to build the ML modeling (Table 1).

**Table 1.**
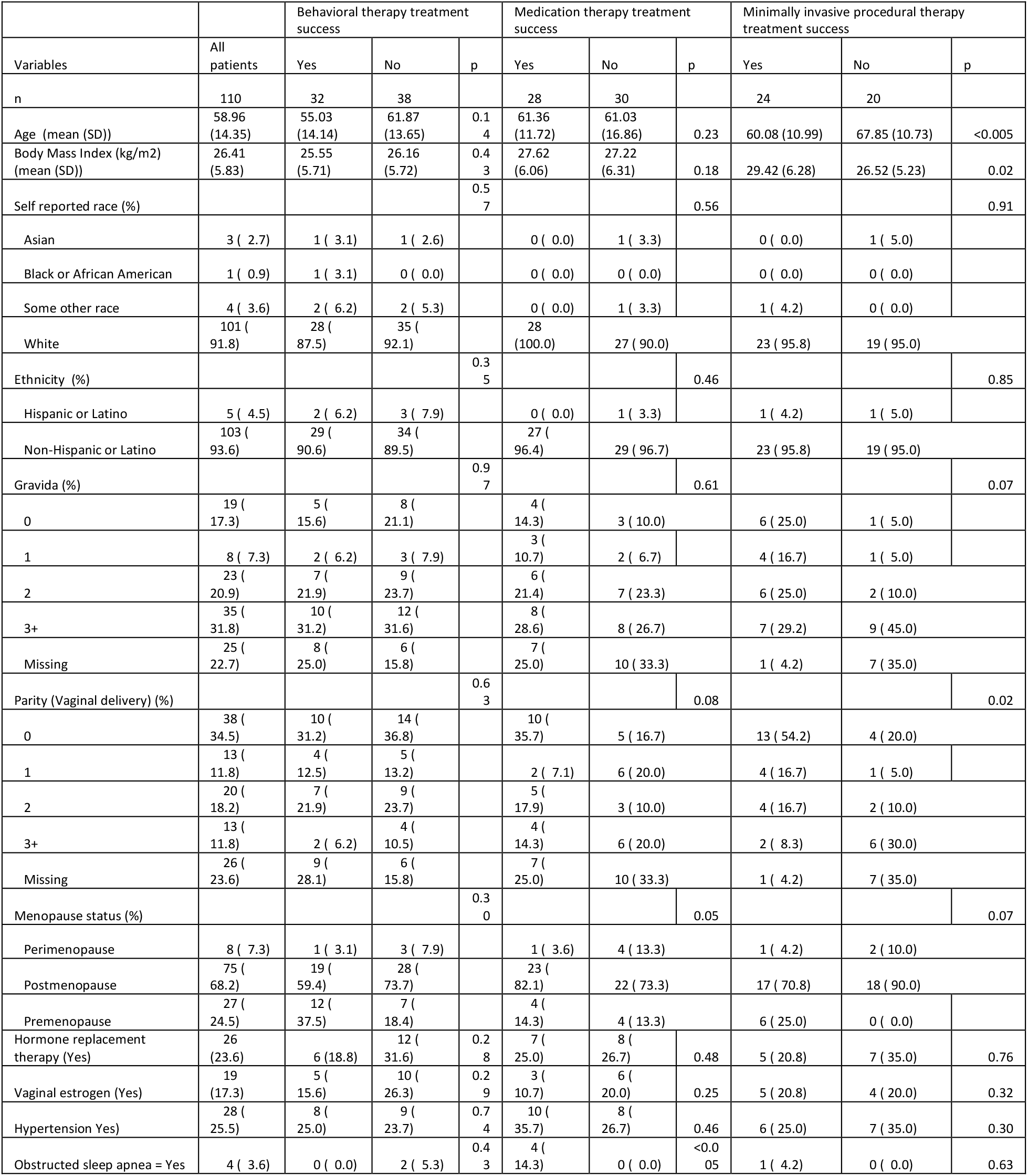
Comparison of patient demographics for those with treatment success stratified by the three treatment subtypes.

**Table 2.** Performance off LASSO logistic and xgboost machine learning modeling for all treatment therapies.

| Treatment therapy | Sensitivity | Specificity | Area under the Curve |
| --- | --- | --- | --- |
| **LASSO logistic regression** |  |  |  |
| Behavioral therapy | 0.45 | 0.69 | 0.49 |
| Medication therapy | 0.48 | 0.77 | 0.76 |
| Minimally invasive procedural therapies | 0.65 | 0.78 | 0.83 |
| **xgboost** |  |  |  |
| Behavioral therapy | 0.45 | 0.69 | 0.49 |
| Medication therapy | 0.60 | 0.65 | 0.74 |
| Minimally invasive procedural therapies | 0.56 | 0.46 | 0.6 |

For the behavioral therapies, feature selection resulted in the following top-ranking inputs: age, hormone replacement therapy, prior hysterectomy, use of fumarates for MS treatment, subjective urinary retention/sensation of incomplete bladder emptying, presence of pelvic organ prolapse, detrusor underactivity, no SUI on UDS, bladder capacity and MS lesion in the deep white matter of brain MRI. The LASSO logistic regression modeling had a sensitivity of 0.68 and specificity of 0.62 with an AUC of 0.74 (Figure 1).

**Figure 1.**
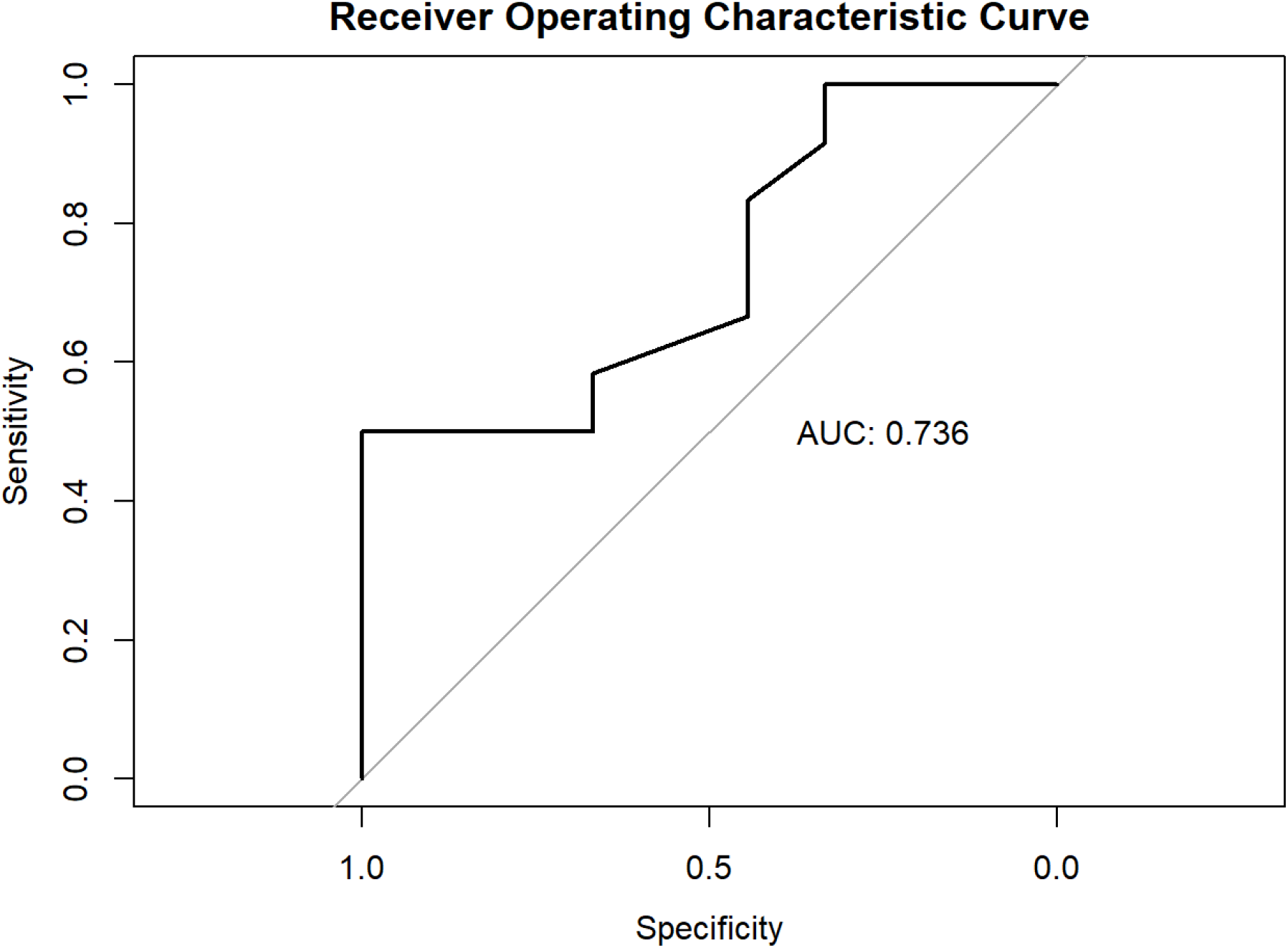
Receiver operating characteristic curve using LASSO logistic regression to predict treatment success with behavioral therapy.

A non-linear prediction model xgboost was created but the AUC was 0.49. On the SHAP plot, which evaluates the average influence of the variable on model prediction, no SUI on UDS and subjective reports of incomplete bladder emptying had the highest impact on model prediction.

For medication therapies, feature selection resulted in the following top-ranking inputs: age, obstructed sleep apnea, prior hysterectomy, subjective sense of urinary retention, bladder capacity and detrusor overactivity on urodynamics and MS lesions on the posterior spinal cord and MS lesions in the supratentorial, infratentorial and deep white matter of the brain. On LASSO logistic regression, the sensitivity was 0.48 with specificity of 0.78 with an AUC of 0.76 (Figure 2). On non-linear predictive model xgboost, the AUC was 0.74. On SHAP plot, subjective sense of incomplete bladder emptying, increased bladder capacity on urodynamics and presence of MS lesions in the deep white matter of the brain and posterior spinal cord had the highest impact on the final model prediction.

**Figure 2.**
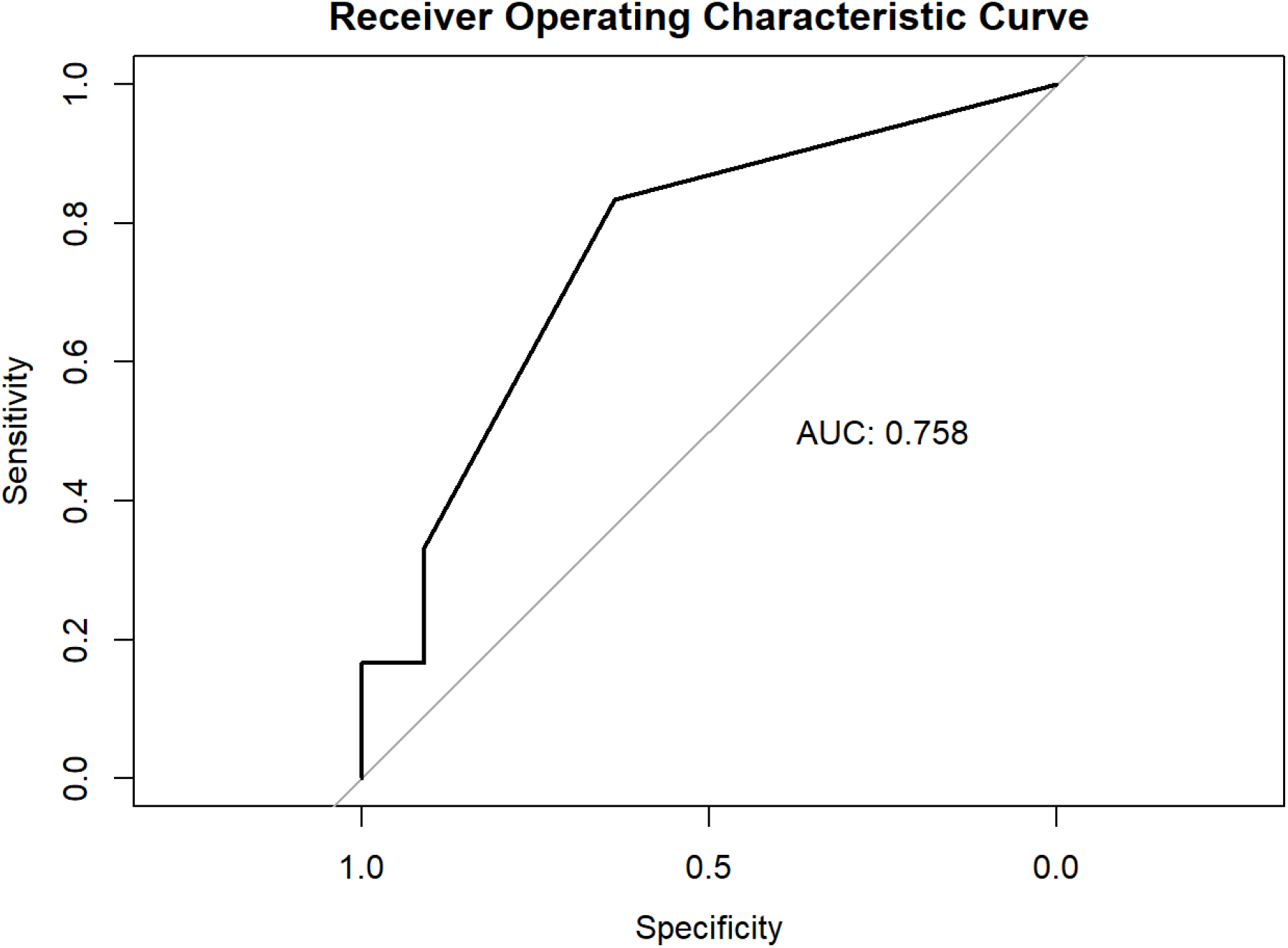
Receiver operating characteristic curve using LASSO logistic to predict treatment success with medication therapy.

For minimally invasive therapies, feature selection resulted in the following top-ranking inputs: age, gravida, menopause status, active medication management for MS, objective incomplete bladder emptying, bladder capacity and findings of stress urinary incontinence, detrusor overactivity and absence of Valsalva voiding on urodynamics, and MS lesion along thoracic spine. On LASSO logistic regression, the sensitivity was 0.65 with a specificity of 0.78. The AUC with LASSO logistic was 0.83 (Figure 3). On non-linear prediction modeling with xgboost, the AUC was 0.6. On SHAP plot, the features that contributed most to the model included increasing bladder capacity, absence of Valsalva voiding and detrusor activity on urodynamics as well as increasing age.

**Figure 3.**
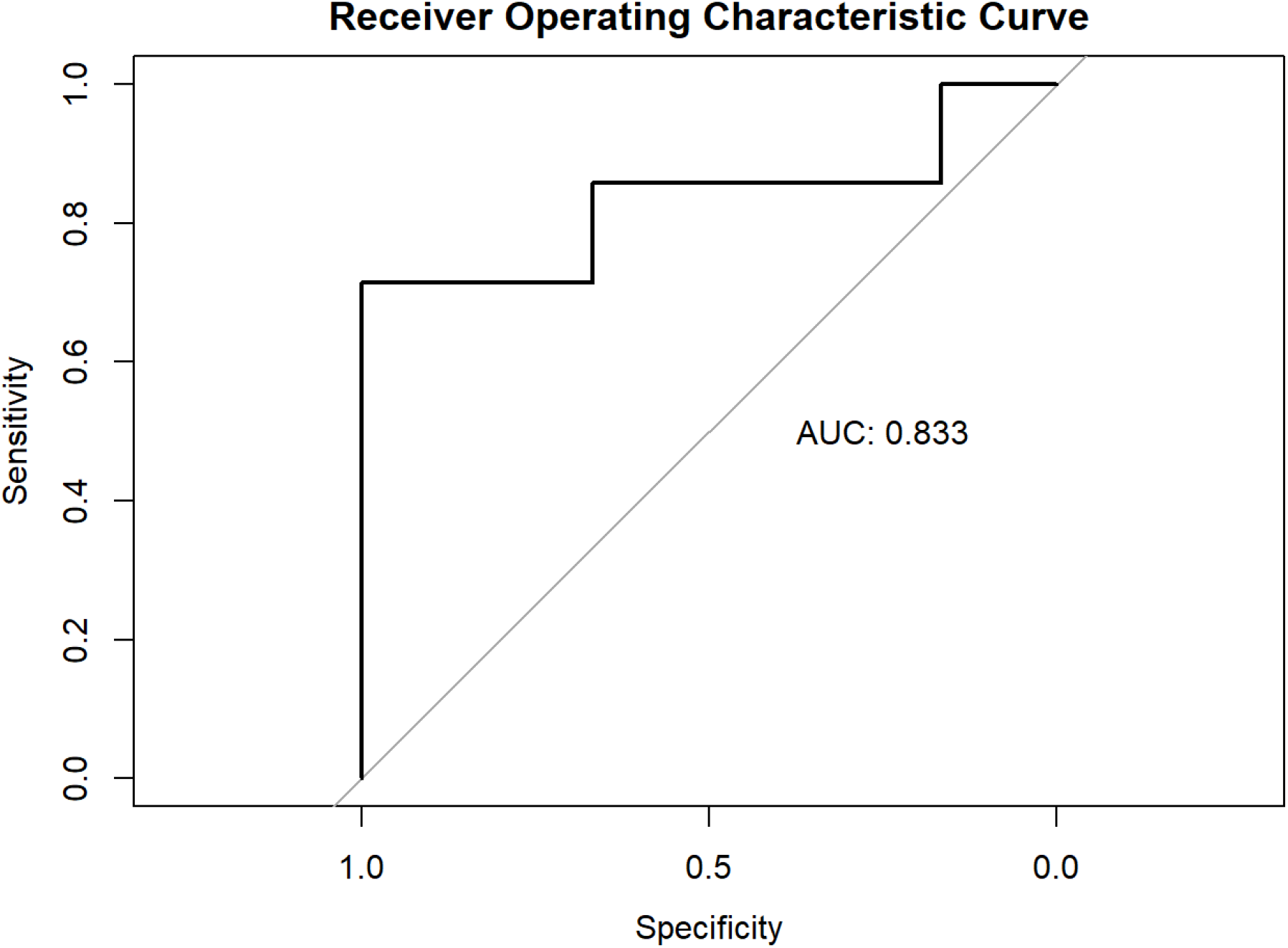
Receiver operating characteristic curve using LASSO logistic to predict treatment success with surgical therapy.

## Discussion

With various machine learning models aimed at predicting treatment success for various types of treatment for neurogenic overactive bladder, we successfully identified several predictors of treatment success including concurrent medical diagnoses, certain urodynamic findings and presence of MRI lesions along the spinal cord and brain.

The economic burden of urinary dysfunction is significant with a cost of $65.9 billion dollars annually in the United States.^13^ In patients living with both NLUTD and MS, there is dual impact on quality of life. This underscores in the importance of increasing access to care and minimizing the time need to establish high quality care. Several machine learning models have been developed in predicting objective and patient-reported outcomes for OAB treatment utilizing various datasets.^14,15^ These ML models have high accuracy with AUCs as high as 0.95 for predicting treatment success with chemodenervation of the bladder.^15^ These studies, along with our findings, highlight the importance of further developing and refining ML models to optimize treatment prediction based on individual patient factors.

On ML modeling for behavioral therapies, absence of SUI on UDS and sensation of incomplete bladder emptying had the highest impact on model prediction. Many patients with OAB report sensation of incomplete bladder emptying; when asked, this is described as frequent voids with a self-perception of an incompletely empty bladder rather than an obstructive voiding pattern from bladder outlet obstruction. Broadly speaking, the SHAP ML model suggests that traditional behavioral therapy such as fluid management and bladder retraining may work best for those with subjective perception of urinary dysfunction, and for those who are “dry” without urinary incontinence.

Interestingly, presence of MS lesions along the brain and spinal cord had the highest impact on ML modeling for treatment success with medication therapies, including anticholinergic and beta-3-agonists. Studies have shown that areas in the brain with higher mean diffusivity with evidence of axonal degeneration and demyelination as seen in MS lesions are associated with higher urinary distress on validated questionnaires.^16^ In another cohort of MS patients undergoing functional MRI, it was found that 2 additional white matter tracts were associated with voiding dysfunction,^17^ and have decreased levels of activation in these key regions of interest regarding urinary urgency and voiding.^18,19^ In one pilot study, there were differences in regional brain activation in those treated with anticholinergic and non-anticholinergic medications.^20^ With the retrospective study design, we were not able to identify the exact location of the MS plaques on MRI reports and correlate that with the medication used. Nonetheless, the modeling created from this preliminary study suggests that MS plaque lesions may play a role in predictive treatment success particularly as it pertains to medication therapy.

Furthermore, from an implementation science perspective, we know that there are effective therapies for idiopathic OAB but adherence is a concern.^21^ Medical treatments include antimuscarinic medications or beta-3-agonists. While antimuscarinic medications are effective, it has bothersome side effects like dry mouth and constipation to concerning potential for dementia over the long term.^22^ Thus, antimuscarinics may exacerbate the known cognitive impairment associated with multiple sclerosis.^23^ Machine learning can allow us to develop patient-specific recommendations with personalized calculators to predict treatment for both OAB and NOAB with a high likelihood of success, rather than trialing traditional step-wise therapies that may be required by insurance companies.

While these ML models showed promising results, there are some limitations, including its retrospective design, small sample size and validation in the same cohort. The current models have high AUCs but would benefit from further expansion of modeling before employing for clinical use. Further studies would benefit from a prospective design to abstract granular MRI data with standardized clinical definitions to ensure accurate and reliable interpretation of clinical data. We acknowledge that this study only captures a subset of NLUTD and the modeling does not include those with concurrent urinary dysfunction. However, this type of exploratory modeling is important to demonstrate that there are potential features to be explored prior to larger, prospective trials.

## Conclusion

Utilizing the ten features of statistical and clinical importance, we constructed various machine learning modeling stratified by the various treatment types of NOAB. With this strategy, we were able to achieve models with predictive value of 0.74-0.83 in its ability to predict treatment success based on individual factors. However, retrospective data is limited by its data input so prospective studies are needed to carefully examine these identified associations.

## Data Availability

The data that support the findings of this study are available from the corresponding author, OC, upon reasonable request.

